# Climate change is already reshaping schistosomiasis transmission across Africa

**DOI:** 10.64898/2026.06.01.26354594

**Authors:** Meghan Forstchen, Ibrahim Aslan, Caroline Bice, Heather Buelow, Andrew J. Chamberlin, Giulio A. De Leo, Kristie L. Ebi, Nicholas A. Galle, Patrick Heffernan, Karena H. Nguyen, Matthew Sisk, Jason R. Rohr

## Abstract

Climate change is shifting infectious disease burdens^1–6^, but attributing transmission changes remains difficult where interventions and socioeconomic development interact with temperature-dependent signals^7–11^. Mechanistic models can isolate temperature-dependent signals from non-climatic influences^5,12–16^ but are often not tested against independent data. Here, we present a validation-first framework using a temperature-dependent R₀ transmission model^17^ to detect and attribute temperature-mediated climate impacts on schistosomiasis transmission across Africa. First, semi-natural mesocosm experiments confirmed the model’s biological constraints, with high temperatures suppressing the host-parasite system above ∼33°C. Next, we established epidemiological relevance in the Lake Victoria Basin using 141,829 longitudinal infection records. Interannual temperature anomalies predicted infection risk, with anthropogenic warming accounting for 17.1% of observed infections relative to a natural-forcing-only counterfactual. Finally, across Africa, the mechanistic R₀ predictor explained prevalence better than correlative climate metrics, even after accounting for intervention and socioeconomic covariates. Applying the validated framework to ensemble climate model simulations and a natural-forcing-only counterfactual (1984–2014) showed that anthropogenic warming increased transmission potential in cooler regions while suppressing it in hotter regions across Africa, a contrast projected to intensify under higher-emissions scenarios by mid-century. Climate impacts are not solely future threats, but present-day forces already reshaping transmission and disease burden.

## Introduction

Climate change is reshaping the global distribution of infectious disease^1–6^; yet aside from a few notable exceptions^18,19^, the extent to which anthropogenic warming has already altered transmission and disease burden remains poorly quantified. While a growing body of research projects future range shifts and climate-driven changes in transmission risk^20–22^, far fewer studies have quantified how much observed change is attributable to anthropogenic warming rather than to non-climatic drivers^18,23–25^. Attribution analysis is notably difficult for neglected tropical diseases (NTDs), where warming-driven changes in transmission risk can be obscured by simultaneous public health interventions and socioeconomic development^7–11^. Consequently, even in systems with well-established thermal sensitivity, stable or declining disease burden can blur the contribution of temperature-driven changes to transmission risk^7–11^.

The NTD schistosomiasis provides a powerful test case for this attribution challenge. Second only to malaria in global parasitic disease burden, it affects more than 230 million people globally^26–28^, with most cases concentrated in children^27^ residing in Sub-Saharan Africa^28^. Its transmission depends on obligate freshwater snail hosts and free-living aquatic parasite stages^29,30^, and is constrained by nonlinear thermal responses in both host and parasite physiology^5,17,31,32^. Anthropogenic warming should therefore reorganize transmission risk unevenly across space, perhaps by expanding suitability in historically cool regions while suppressing it where temperatures exceed physiological optima^9,33–37^. Crucially, these ecological shifts occur against a backdrop of control measures, particularly mass drug administration (MDA) and water access and sanitation infrastructure (WASH), as well as broader socioeconomic development, all of which can reduce observed disease burden^28,38–40^.

Mechanistic transmission models offer a way to address this detection-and-attribution (D&A) challenge by embedding temperature-sensitive physiological constraints into estimates of transmission potential^5,12–16^. In D&A, detection requires identifying changes in transmission-relevant thermal conditions or temperature-dependent transmission potential, while attribution compares those changes against a counterfactual climate baseline without anthropogenic forcing. However, D&A is only credible if the underlying predictive model is validated against independent evidence across relevant biological, epidemiological, and geographic scales^5,41,42^. Without such validation, it remains unclear whether the modeled temperature dependence captures the real-world transmission dynamics and local thermal limits needed for credible attribution and projection.

Here, we adopt a validation-first framework to detect and attribute the historical contribution of anthropogenic warming to schistosomiasis transmission and to project future changes in transmission potential under continued warming in Africa. Our work builds on the reproduction number (R₀) model introduced by Aslan et al.^17^ for intestinal schistosomiasis caused by *Schistosoma mansoni* in Africa. By integrating literature-derived thermal responses across the parasite’s free-living and intermediate snail stages, this framework estimates R₀ as a function of temperature to provide a mechanistic measure of environmental transmission potential. Because warming can either amplify or suppress transmission without necessarily crossing a binary suitability threshold, we treat continuous change in model-derived R₀ as the primary endpoint for detecting temperature-dependent shifts in transmission potential.

To assess the model’s predictive power, we first test the mechanistic framework against three independent lines of evidence (Fig. 1a). We begin by validating the biological temperature dependence of transmission using semi-natural, long-term mesocosm experiments that quantify snail dynamics and cercarial production. We then establish its epidemiological relevance using 141,829 longitudinal observations of human infection from the hyper-endemic Lake Victoria Basin^43^. By applying an econometric Mundlak model to isolate interannual temperature anomalies from time-invariant community differences, we show that temperature-dependent signals are detectable in longitudinal infection prevalence despite low spatial variance and active MDA^39^. Finally, we assess spatial generality by testing whether the full mechanistic framework explains continental-scale prevalence patterns across Africa, and whether it improves on standard correlative climate metrics after accounting for strong gradients in socioeconomic development and intervention history. Only after completing these validation steps, do we use the model (Fig. 1b) to attribute historical changes in transmission potential to anthropogenic warming and project future changes under continued warming across Africa (Fig. 1c).

**Figure 1.**
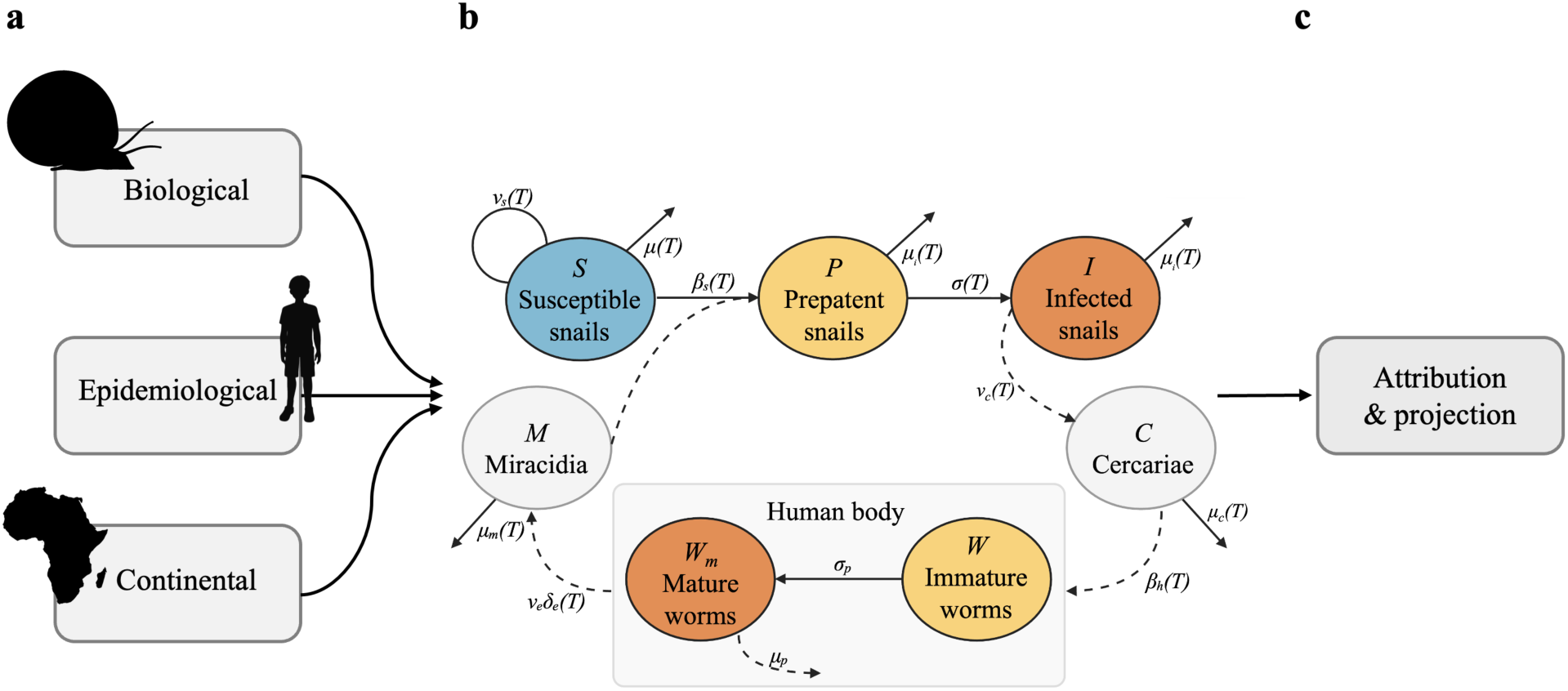
A validation-first framework for attributing anthropogenic-warming impacts on schistosomiasis transmission. (a) The validation-first framework that evaluates the mechanistic transmission model across three scales: biological realism (via semi-natural mesocosm experiments), local epidemiological relevance (via longitudinal infection dynamics and attribution in the Lake Victoria Basin), and continental spatial generality (via geostatistical prevalence modeling). (b) The schematic of the temperature-dependent *Schistosoma mansoni* reproduction number (R₀) model from Aslan et al., (2024). The compartmental framework captures the parasite life cycle across intermediate snail hosts (Susceptible, Prepatent, and Infectious), free-living environmental stages (Miracidia and Cercariae), and the human host (Immature and Mature worms). (c) Application of the validated framework to continental anthropogenic-warming attribution and SSP-based future projections.

### Experimental validation of thermal constraints

To establish the biological realism of the mechanistic framework, we first evaluated whether its predicted temperature dependence of schistosomiasis transmission is upheld under semi-natural conditions (Extended Data Fig. 1). Using freshwater mesocosms spanning a realistic thermal gradient for schistosomiasis transmission in Africa (18–37°C), we tracked *Biomphalaria glabrata* host population dynamics and the production of *S. mansoni* cercariae, the parasite stage infectious to humans, over 16 weeks. Snail populations thrived at intermediate temperatures but declined at high temperatures, while cercarial production exhibited a narrower thermal window and was sharply suppressed above ∼33 °C, even where snails persisted, decoupling host presence from transmission potential (Fig. 2; Extended Data Fig. 2; Extended Data Table 1).

**Figure 2.**
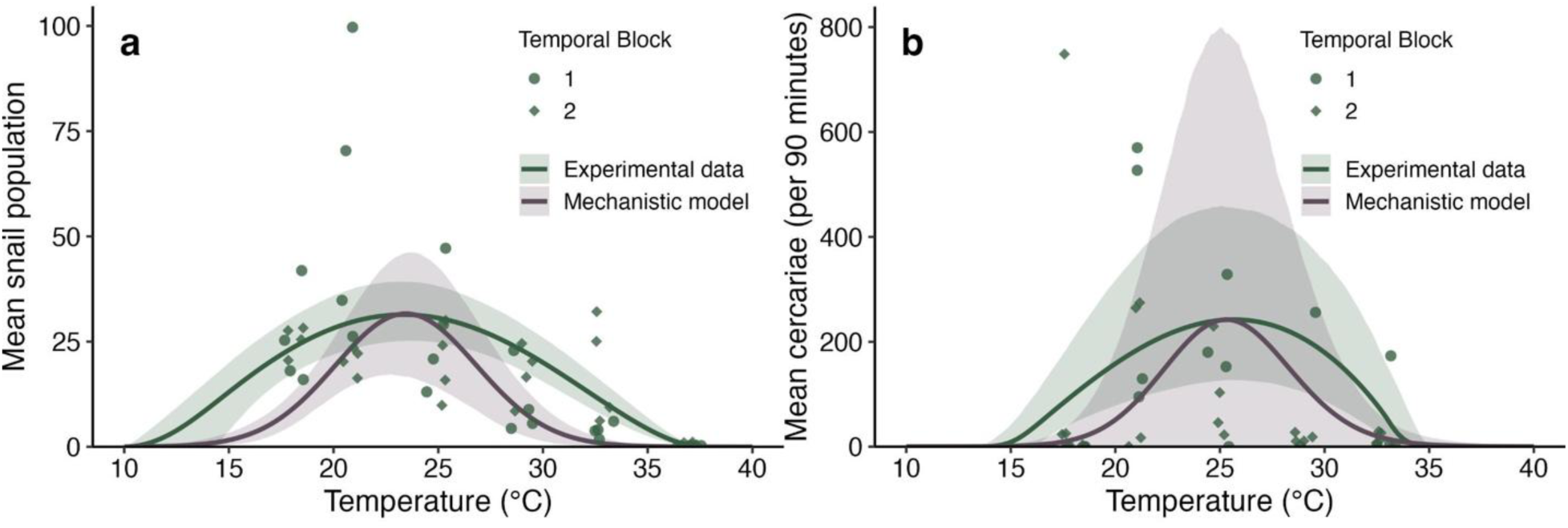
Comparison of experimental and mechanistic thermal performance curves. The comparison of experimentally derived and mechanistically derived (from the Aslan et al. model) thermal performance curves (TPCs) for (a) mean snail population and (b) mean cercarial production. Green lines and ribbons represent Bayesian TPCs (mean and 95% credible intervals) fitted to experimental data. Points represent observed values for temporal blocks 1 (circles) and 2 (diamonds). Purple lines and ribbons represent TPCs (mean and 95% confidence intervals) derived from a two-step ensemble of the mechanistic transmission model and bootstrapped to match experimental sample sizes.

The mechanistic model captured the overall humped shape and key thermal features of the experimentally derived thermal performance curves (TPCs), including the location of thermal optima and the sharp decline in transmission at high temperatures (Fig. 2, Extended Data Table 1). Although the model underestimated the upper thermal limit for snail survival relative to our mesocosm observations, this discrepancy rendered the model conservative, and it predicted high-temperature persistence and the cessation of transmission in the low 30s (°C). Together, these results provide the first out-of-sample experimental validation of the mechanistic transmission framework, showing that its temperature dependence reflects realized biological constraints rather than artifacts of model fitting.

### Epidemiological detection and attribution in the Lake Victoria Basin

Next, to demonstrate epidemiological relevance, we assessed whether the predicted temperature dependence is detectable in human infection outcomes, while accounting for intensive MDA^39,44^, precipitation, distance to the lake, and water access. Using 141,829 longitudinal observations of *S. mansoni* infections in school-age children across 365 lakeshore communities (SCORE study villages) in the Lake Victoria Basin from 2011-2015^39,44^, we fit a beta-binomial generalized linear mixed model (GLMM) with a Mundlak specification to test whether the predicted temperature dependence is detectable in a major schistosomiasis transmission hotspot^43^. This specification separates year-to-year temperature deviations within each community from stable differences across communities. It therefore isolates interannual temperature effects, while accounting for unobserved community differences that remain relatively stable over the study period, such as baseline ecology, human behavior, and health system characteristics.

Across the observed temperature range in the study area (Extended Data Fig. 3), which fell largely on the ascending portion of the mechanistic R₀ curve, model-predicted R₀ was nearly perfectly correlated with annual temperature (*r* = 0.991). We therefore used within-community temperature anomalies, defined relative to each community’s mean over the study period, rather than model-derived R₀, as the predictor of transmission risk. The Mundlak model revealed that positive within-community temperature deviations were significantly associated with increased *S. mansoni* prevalence (*β_within_* = 0.14, p=0.004; Fig. 3a). These associations persisted after accounting for MDA, precipitation anomalies, and village-level random effects (full model Marginal R^2^ = 0.460), which provides strong epidemiological evidence that interannual warming has amplified human infection risk in this region of Africa (Extended Data Table 2).

**Figure 3.**
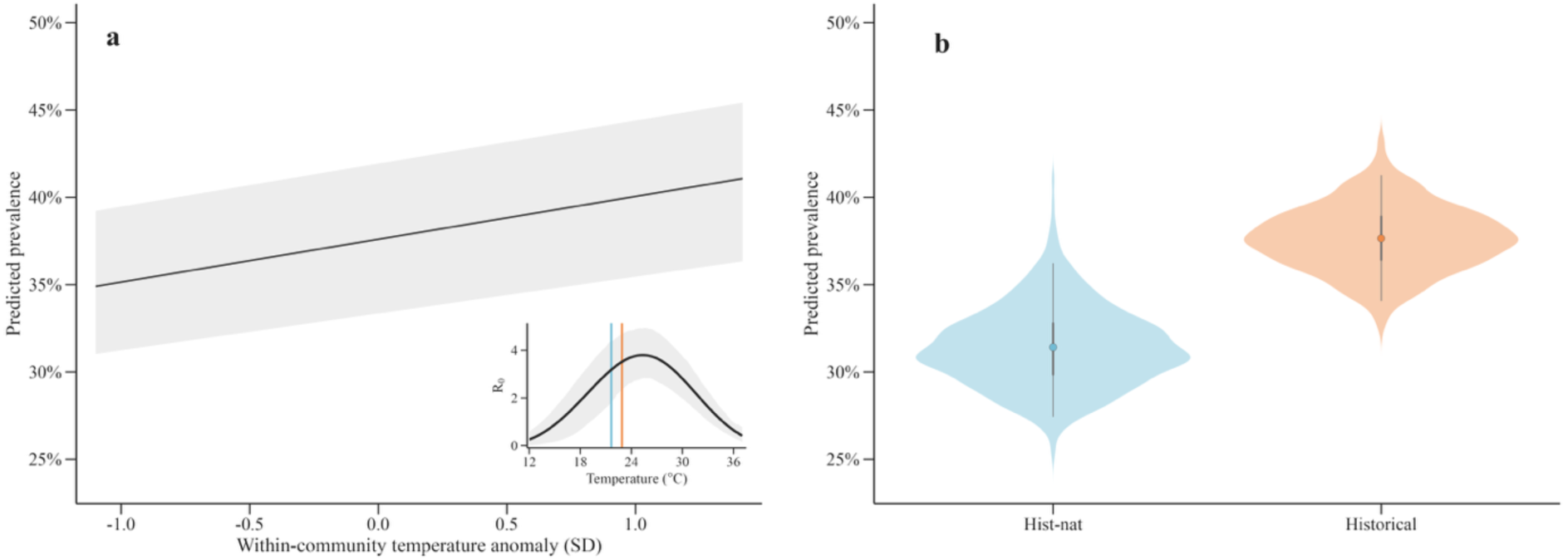
Anthropogenic warming increases predicted *S. mansoni* prevalence around Lake Victoria. (a) Model-adjusted predicted prevalence from SCORE surveys in communities within 10 km of Lake Victoria, plotted against within-community temperature anomalies. The line shows the fitted marginal trend from a Mundlak beta-binomial model averaged over observed covariates; shading indicates the 95% uncertainty interval. The inset shows the temperature-dependent *S. mansoni* R₀ curve, with the blue and orange vertical lines marking Lake Victoria temperatures under natural-forcing-only (hist-nat) and factual historical climate conditions, respectively. (b) Counterfactual predicted prevalence under natural-forcing-only and factual historical climate conditions. Violin plots show simulated predicted-prevalence distributions; points indicate mean predictions and vertical bars show 50% and 95% intervals. Analyses included 1,285 community-year observations from 365 communities, representing 141,829 children examined. Historical climate increased predicted prevalence by 6.1 percentage points, corresponding to an attributable fraction of 17.1% (95% CI, 6.0–23.4%) when calculated from simulated, sample-size-weighted expected infections rather than from rounded plotted prevalence summaries.

To quantify the burden attributable to anthropogenic warming, we predicted school-age infection prevalence in each community under a natural-forcing-only counterfactual temperature series with the anthropogenic signal removed, using historical and natural-forcing-only climate model ensembles from the Coupled Model Intercomparison Project Phase 6 (CMIP6)^45^. Using the estimated within-community temperature effect on infection risk (*β_within_*), this counterfactual cooling reduced mean predicted prevalence from 36.4% to 30.3%, an absolute difference of 6.1%; when calculated from simulated, sample-size-weighted expected infections, this corresponded to an attributable fraction of 17.1% (95% CI, 6.0–23.4%) of observed infections in the longitudinal dataset (Figure 3b). Assuming this attributable fraction scales to the total ∼256,600 km^2^ Lake Victoria Basin (Extended Data Fig. 3), we estimate that anthropogenic warming contributed 382,539 excess cases (95% CI: 146,260–560,913) and 3,777 additional DALYs (95% CI: 1,263–6,018) among children aged 5–14 in 2015 alone.

### Continental validation of mechanistic transmission constraints

Demonstrating mechanistic consistency in a single region does not establish generality across Africa’s heterogeneous ecological, climatic, and intervention landscapes. We therefore evaluated whether the mechanistic framework explains broad spatial patterns of schistosomiasis prevalence at the continental scale. Because repeated, community-level sampling was unavailable at the continental scale, longitudinal attribution was not possible. We therefore evaluated spatial validity using 14,643 *S. mansoni* prevalence estimates from 697,919 individuals across 21 African countries (2015–2023)^46^ by fitting geostatistical models. We compared a baseline model of non-thermal environmental, socioeconomic, and intervention covariates (Supplementary Table 1) against models that incorporated either: raw temperature metrics, experimentally derived TPCs for snail population or cercarial production, mechanistic model-derived TPCs for the same traits, or the integrated temperature-dependent mechanistic R₀ from Aslan et al.^17^.

The results highlight the value of biological mechanism over statistical correlation alone. Adding raw temperature metrics (T_min_ and T_max_) to the baseline model yielded only marginal gains in predictive performance (ΔAUC = +0.017), whereas biologically derived thermal predictors consistently improved model performance. Models incorporating experimental or mechanistic TPCs outperformed the raw-temperature model, and the experimental population TPC and integrated temperature-dependent R₀ model jointly achieved the largest gain in predictive accuracy (ΔAUC = +0.054; mean AUC = 0.849; Extended Data Table 3). These patterns persisted under spatially structured partitioning, with the R₀ model retaining strong predictive performance and overlapping AIC support with the highest-AUC model under spatial checkerboard cross-validation (Supplementary Table 2).

Analysis of the mean permutation importance (PI) of predictors demonstrates why temperature-dependent climate signals are often obscured in clinical data. In the best performing model, MDA coverage was the single strongest predictor of prevalence (PI: 0.162), followed by GDP (0.131), temperature-dependent R₀ (0.119), elevation (0.105), and water access (0.060). Collectively, these factors accounted for more than half of the increase in predictive accuracy of prevalence (Extended Data Table 3). These results indicate that temperature-dependent transmission constraints help explain prevalence patterns alongside major intervention and socioeconomic predictors, providing a third out-of-sample test of the mechanistic framework, this time at the continental level.

### Historical attribution and future projections of transmission potential

After validating the mechanistic framework across biological, epidemiological, and spatial scales, we quantified the magnitude of anthropogenic warming signal by comparing climate model simulations with and without human forcing (CMIP6 historical vs natural-forcing-only) over 1984–2014. Across Africa, anthropogenic warming increased temperatures by a mean of 0.92 °C (±0.24 °C), with positive anomalies in all land grid cells (2.5th–97.5th percentile: +0.63 to +1.18 °C; Extended Data Fig. 4).

We then attributed the effects of this anthropogenic warming on transmission potential by propagating this signal through the validated R₀ model. For each grid cell and year from 1984–2014, we calculated R₀ under CMIP6 historical temperatures (R₀,_historical_) and under the natural-forcing-only counterfactual (R₀,_hist-nat_), defining the anthropogenic transmission effect as ΔR₀ = R₀,_historical_ − R₀,_hist-nat_. Inter-model variability and sign agreement for this propagated ΔR₀ signal are shown in Extended Data Fig. 5.

This comparison revealed that anthropogenic warming has already increased transmission potential in cooler regions and suppressed it in hotter regions across Africa (Figs. 4a, 4b, 5a). In historically hot Sahel (e.g., Senegal, Mali, and South Sudan), warming reduced transmission by exceeding physiological optima, whereas in cooler East African highlands and southern subtropical regions (e.g., Rwanda, Zambia, and Madagascar), it increased transmission potential by relaxing thermal constraints. Overall, approximately 611 million people reside in areas where warming amplified transmission potential, compared with approximately 410 million in areas of suppression (Fig. 5a).

**Figure 4.**
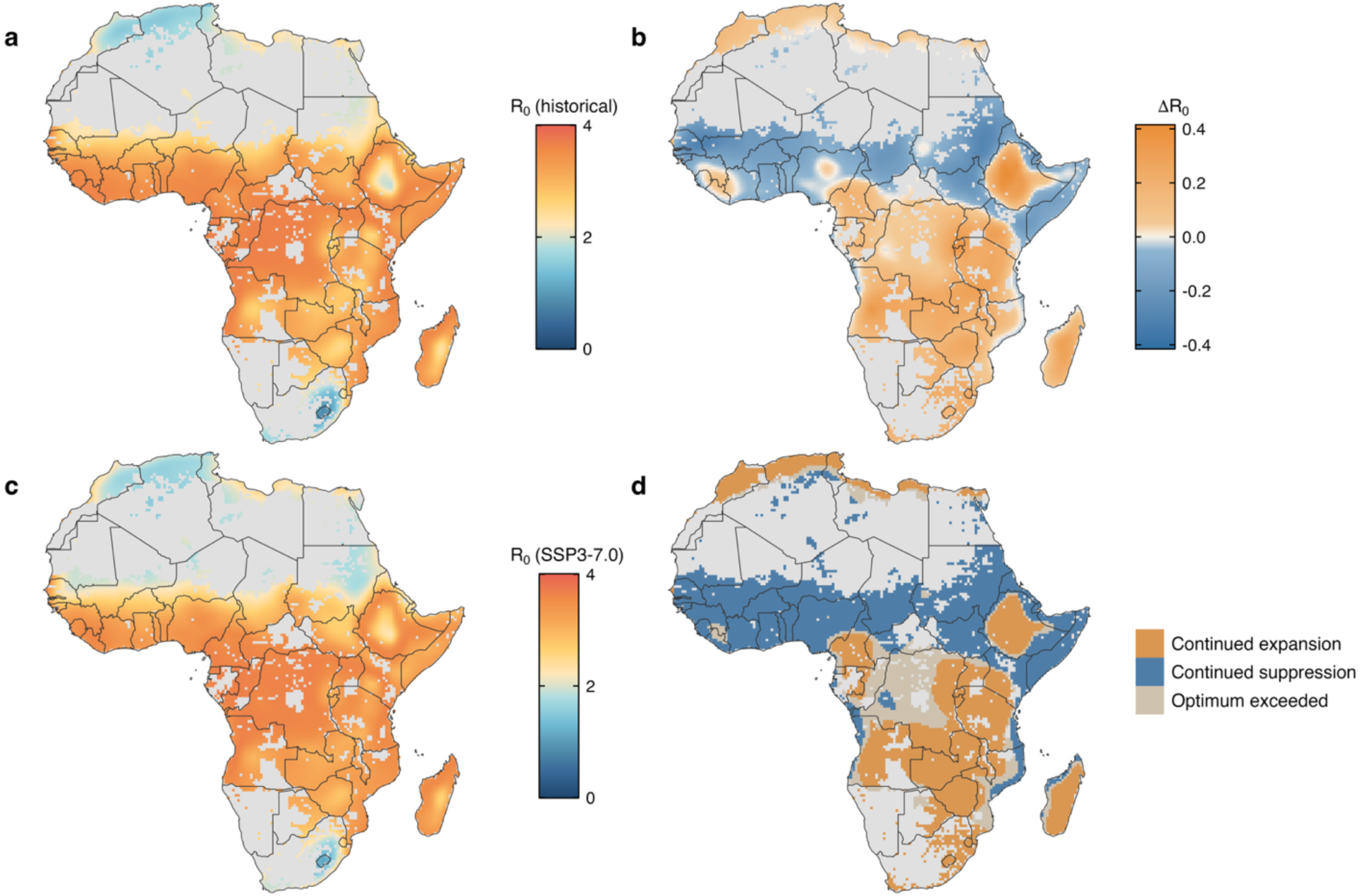
Historical attribution and future trajectories of schistosomiasis transmission potential across Africa. (a) Mean historical transmission potential (R₀) for *S. mansoni* from 1984-2014, derived from the validated mechanistic model. (b) The attribution to anthropogenic warming, calculated as the *Δ*R₀ between the factual historical record and a natural-forcing counterfactual scenario. Orange indicates areas where anthropogenic warming has already amplified transmission potential, while blue indicates areas of thermal suppression where temperatures have moved beyond the physiological optimum. (c) Projected mean transmission potential by mid-century (2020-2050) under the SSP3-7.0 emissions scenario. (d) Qualitative classification of transmission trajectories. ‘Continued expansion’ (orange) denotes regions where transmission potential has increased historically and is projected to rise further. ‘Continued suppression’ (blue) indicates historically suppressed areas that will experience further declines in transmission potential. ‘Optimum exceeded’ (tan) highlights transition zones where historical warming initially expanded the transmission window, but future warming is projected to exceed physiological limits and trigger thermal suppression.

**Figure 5.**
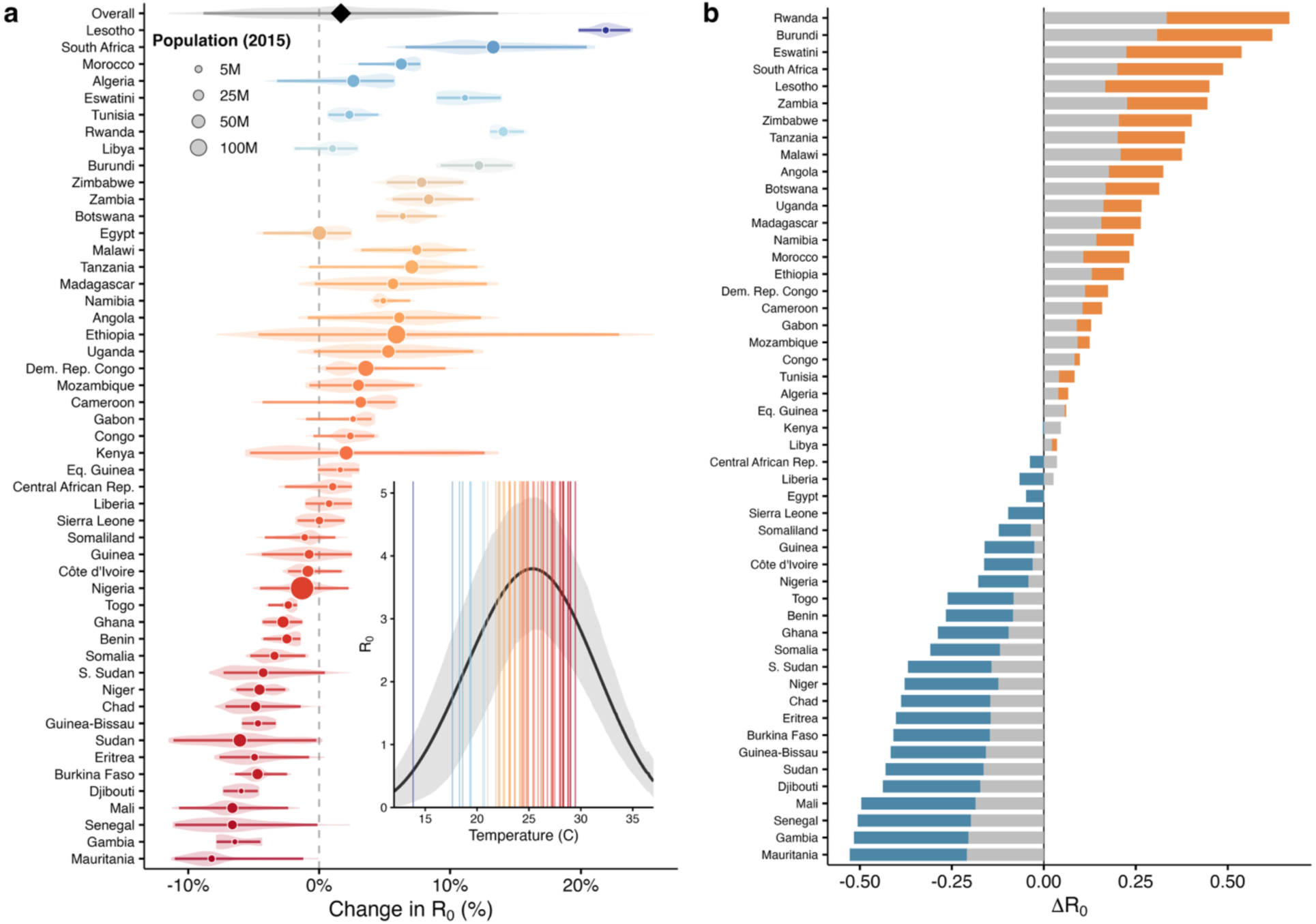
Change in schistosomiasis transmission potential. (a) change in R₀ (%) calculated by comparing the historical temperature against a natural-forcing counterfactual baseline (1984-2014). Points represent the mean proportional change for each country, with point sizes scaled to the national population (2015). Shaded horizontal intervals and violins represent the 2.5th and 97.5th percentiles of spatial heterogeneity in transmission shifts within each country. (b) Comparison of historical climate attribution (1984-2014) and projected mid-century changes (2020-2050) in *S. mansoni* transmission potential (*Δ*R₀). Grey bars represent the magnitude of change already attributable to anthropogenic warming relative to a natural-forcing counterfactual. Colored bars indicate the additional projected change under the high-emissions SSP3-7.0 scenario. Orange denotes projected increases in transmission potential (continued expansion), while blue denotes projected decreases (future thermal suppression).

Under mid-century climate projections (2020–2050) from CMIP6 models under a higher-emissions scenario (SSP3–7.0), the mechanistic model indicates that these historical trends will strengthen (see Extended Data Fig. 6 for other scenarios). Thermal suppression of transmission potential is projected to intensify across much of West Africa, while transmission potential is projected to increase further in the East African highlands and southern subtropics (Figs. 4c & 4d). In parts of the central African lowlands, warming is projected to shift conditions from increasing to suppressing transmission as temperatures exceed physiological optima (Figs. 4c & 4d). Comparing the historical period (1984–2014) with these projections shows that 44.5% (95% CI: 42.6–46.5) of the total mid-century shift has already occurred by 2014 (Fig. 5b). These results indicate that climate impacts are not solely future threats, but ongoing processes already reshaping transmission landscapes.

## Conclusions

Although climate change is widely recognized as a growing challenge for infectious disease control, our results show that its ecological and epidemiological impacts on schistosomiasis are already detectable and substantial. Using a validation-first framework, we show that anthropogenic warming has altered transmission potential, increased human infections, and contributed to disease burden in Africa, even in the presence of widespread control efforts.

A central implication is that stable or declining prevalence does not imply climatic irrelevance. Public health interventions can reduce overall burden while temperature-driven transmission pressure increases, thereby masking the climate-related component of observed disease trends. As a result, the proportion of remaining burden attributable to anthropogenic warming may increase even as total prevalence declines. This complicates interpretation of epidemiological trends and underscores the need for mechanistic climate attribution approaches that distinguish intervention-suppressed burden from reduced transmission potential, which is critical for identifying areas where control gains may be fragile under continued warming.

By integrating experimental biology, local epidemiology and continental-scale spatial analysis, we provide a strong basis for attributing climate impacts on schistosomiasis transmission. The concordance across independent biological, epidemiological and spatial analyses suggests that temperature-dependent transmission constraints are a general feature of *S. mansoni* transmission biology in Africa, rather than an artifact of any single dataset or modelling approach. This framework offers a generalizable template for attributing climate impacts in disease systems where interventions, socioeconomic change and environmental change co-occur.

More broadly, these findings show that climate attribution in infectious disease systems is feasible even where intervention and socioeconomic change obscure environmental signals. Disease control can no longer assume that present-day transmission reflects a stable climatic baseline. Instead, strategies must be recalibrated for a climate-altered present rather than deferred to an uncertain future.

## Methods

### Mesocosm experiments

To evaluate the mechanistic model compartments under semi-natural conditions, we conducted a 16-week mesocosm experiment simulating freshwater ecosystems across a thermal gradient. The experiment followed established protocols^47^, using 17-L mesocosms maintained at 6 constant temperature treatments (18, 21, 25, 29, 33, and 37°C; *n*=4 replicates per temperature per block; 2 temporal blocks) (Extended Data Fig. 1).

Mesocosms were inoculated with algal and zooplankton communities collected from local ponds and allowed to establish for 10 days prior to the introduction of uninfected *B. glabrata* snails (NMRI strain; *n*=5 per tank) from the Schistosomiasis Resource Center (SRC) at the Biomedical Research Institute (Rockville, MD). *B. glabrata* was selected because African *Biomphalaria* lines were not available through the SRC and because *B. glabrata* is a widely used laboratory model for *S. mansoni* transmission studies. *S. mansoni* eggs (NMRI strain) harvested from infected mouse livers, provided by the SRC, were introduced at weeks 1, 3, 6, and 9. Egg viability was quantified prior to inoculation (Extended Data Fig. 1c). We monitored snail population dynamics and cercarial production weekly. All snails >2 mm were measured for growth analysis (Extended Data Fig. 1d), and patent infections were quantified by inducing cercarial shedding under light stimulation. Cercarial counts were performed directly or via validated grid-subsampling for high-density samples (Extended Data Fig. 1a). At the experiment’s conclusion, all remaining snails were necropsied to confirm infection status.

Snail population density and total cercarial production were analyzed using Generalized Additive Mixed Models (GAMMs) in R via the package *mgcv*^48^ to capture nonlinear thermal responses over time. Models included temperature and week as tensor product smooths, with experimental block as a fixed effect and tank identity as a random effect. A Tweedie distribution was selected for both response variables based on residual diagnostics and AIC comparison (Extended Data Fig. 2).

### Thermal performance curve comparison

We adapted the *S. mansoni* transmission model^17^ to match the mesocosm conditions and thereby enable direct comparison between theoretical predictions and experimental results. The model structure (susceptible-prepatent-infected *Biomphalaria* spp., miracidia, cercariae) was parameterized with previously published temperature-dependent traits but initialized with the same snail abundances and pulsed egg introduction schedules used in our mesocosms. We simulated dynamics using the *odin*^49^ package for 16 weeks across a 10–40°C range and calculated weekly mean snail abundance and cercarial production to match experimental sampling intervals.

We propagated parameter uncertainty for the transmission model using a two-step ensemble approach. First, we generated 500 model simulations by sampling input parameters from multivariate normal distributions defined by their original estimation covariance. Second, to match the finite sample size of the experiment (*n*=8 independent mesocosm replicates per temperature), we created a mechanistic bootstrap ensemble by resampling these simulations. We then fit parametric (Gaussian) thermal performance curves (TPCs) to these ensembles using non-linear least squares via the *nls.multstart* package^50^ to derive mean trajectories and 95% confidence intervals for snail abundance and cercarial shedding.

For the experimental TPCs, we quantified the thermal response of the experimental data using a Bayesian framework. Aggregated tank-level data for snail abundance and cercarial production were fit to candidate non-linear thermal functions (Quadratic, Brière, and Modified Brière) assuming a Negative Binomial error distribution to account for overdispersion. Models were fit in JAGS via the *R2jags* package^51^ using weakly informative priors selected via sensitivity analysis. Posterior distributions were estimated via MCMC sampling in JAGS using three chains of 25,000 iterations each, with a burn-in of 5,000 iterations and thinning every 8th draw. Candidate functional forms and prior specifications were compared using the Deviance Information Criterion (DIC). For the selected model, convergence was assessed by visual inspection of trace plots, and model adequacy was evaluated using posterior predictive checks comparing observed data with replicated distributions and summary statistics.

We evaluated model accuracy by comparing the mechanistic and experimental TPCs, along with their 95% uncertainty intervals. Key thermal traits (T_opt_, T_min_, T_max_) were derived from both curves using a 5% performance threshold, which allowed for a quantitative assessment of the model’s ability to capture the realized thermal niche of the parasite system (Extended Data Table 1).

### Epidemiological detection and attribution in the Lake Victoria Basin

We examined anthropogenic warming impacts using longitudinal *S. mansoni* infection data from the Schistosomiasis Consortium for Operational Research and Evaluation (SCORE) trial in the Lake Victoria Basin as a case study^39,44^. This SCORE dataset is a large-scale, multi-year longitudinal cluster-randomized trial tracking *S. mansoni* infection dynamics across communities in Kenya and Tanzania. We restricted the analysis to communities within 10 km of Lake Victoria to focus on communities most likely to experience direct lake-associated transmission. This yielded 141,829 parasitological examinations of school-aged children from 365 communities over 2011–2015 (Extended Data Fig. 3). To account for the transmission window prior to sampling and MDA, we linked parasitological surveys to annual temperature and precipitation exposures from the preceding calendar year. Time-varying temperature and precipitation exposures were derived from WorldClim historical monthly weather data, which are CRU TS monthly weather data downscaled and bias-corrected using WorldClim 2.1.

For each community and study year, we estimated the local annual anthropogenic temperature anomaly as the difference between the multi-model means of the CMIP6 historical and hist-nat ensembles at the nearest grid cell, using the CMIP6 processing and ensembling workflow described below in “Climate detection, attribution, and future projections.” Because the epidemiological analysis linked infection outcomes to temperature over the preceding calendar year, this annual anomaly was used to construct a community-year counterfactual exposure. We then applied the anomaly as a delta adjustment to the annual community-level high-resolution temperature estimate derived from the WorldClim historical monthly weather layers^52,53^ to create a counterfactual temperature trajectory that preserved local spatial heterogeneity while removing the ensemble-estimated anthropogenic warming signal.

To quantify the relationship between anthropogenic warming and infection risk, we modeled community prevalence using a beta-binomial generalized linear mixed model (GLMM) with a logit link in the package *glmmTMB*^54^. The model included categorical fixed effects for country and MDA round, which was defined as the number of praziquantel mass drug administration rounds delivered in each community before the parasitological survey. Additionally, the model incorporated standardized continuous covariates for distance to Lake Victoria^55^ and the proportion of the population relying on surface water^56^. To implement a correlated random-effects (Mundlak)^57^ specification, we decomposed the annual temperature and precipitation exposures into (i) within-community interannual deviations from each community’s mean and (ii) between-community long-run means and included both components as fixed effects to distinguish within-community associations from cross-sectional spatial differences. Random intercepts were included for implementation unit and community to account for hierarchical clustering.

We estimated the impact of anthropogenic warming by generating predictions under both factual and counterfactual temperature series across 1,000 parametric simulations. Fixed-effect coefficients were drawn from a multivariate normal distribution defined by the model variance-covariance matrix, and predictions were generated conditional on the estimated random effects. To focus attribution on longitudinal variation within communities, we propagated the counterfactual temperature shift through the within-community temperature coefficient while holding between-community mean components fixed. We then calculated an attributable fraction as the proportional reduction in expected infections under the counterfactual scenario relative to the factual scenario.

To estimate regional impacts, we applied this attributable fraction to baseline disease burden for school-aged children (5–14 years) in the Lake Victoria Basin. Because this analysis was designed to estimate historical burden during the study period rather than project future impacts, we used population and burden estimates matched to 2015. The school-aged population was extracted from 2015 WorldPop data^58^ at 1-km resolution within the boundaries of the Lake Victoria Basin^59^. Baseline regional burden was estimated by combining these demographic data with country-specific prevalence and DALY rates from the Global Burden of Disease (GBD) 2015 study^60,61^. Finally, we integrated uncertainty via Monte Carlo simulation by combining the distribution of attributable fractions with baseline burden draws derived from GBD 95% uncertainty intervals (assuming independence between these uncertainty sources) to obtain 95% uncertainty intervals for attributable cases and DALYs.

### Continental validation of mechanistic transmission constraints

To evaluate the capacity of our mechanistic framework to explain large-scale prevalence patterns, we developed a zero-inflated beta-binomial GLMM using *S. mansoni* prevalence data (2015–2023) from the ESPEN portal. We extracted a suite of environmental, socioeconomic, and intervention-related covariates for each survey site using the *terra* package^62^ in R (Supplementary Table 1). Models were fit using the *glmmTMB*^54^ package in R with Survey Year included as a random intercept. To mitigate multicollinearity, we applied a forward stepwise variance inflation factor (VIF) screen (threshold <10).

We first constructed a “Baseline” model containing only non-thermal predictors. We then constructed a “Climate-Only” reference model by adding raw temperature metrics (T_min_ and T_max_)^52,53^ to the baseline, allowing us to quantify the improvement offered by standard correlative approaches. To test the predictive power of biological mechanism, we generated a suite of “Mechanistic Geostatistical Models” by incorporating the thermally dependent trait values derived from the TPCs as predictors based on the survey location’s minimum and maximum temperature. We tested five distinct biological inputs: (1-2) the experimental TPCs for snail population and cercarial shedding, (3-4) the model-derived TPCs for the same traits, and (5) R₀ derived from the integrated model. Model performance was evaluated using 5-fold cross-validation by comparing the mean discriminatory ability (AUC) and permutation importance across all candidate models.

To assess whether model comparisons were sensitive to spatial autocorrelation, we repeated model evaluation using spatial checkerboard partitioning with approximately 100-km spatial blocks, implemented with get.checkerboard2 in the ENMeval R package^63^. This procedure assigns geographically structured subsets of occurrence locations to alternating spatial bins, providing a more conservative test of predictive performance under spatial non-independence. Models were refit and evaluated across checkerboard partitions, and performance was compared using AUC and AIC. Spatial checkerboard partitioning confirmed that the integrated R₀ model retained strong predictive support, with AIC-based grouping placing it within the same performance class as the top-ranked thermal models (Supplementary Table 2).

### Climate detection, attribution, and future projections

CMIP6 daily near-surface air temperature (tas) data were accessed through the Earth System Grid Federation^45,64^. These data were used to detect and attribute historical changes in temperature-dependent schistosomiasis transmission potential and to project future changes under continued warming. We analyzed six experiments: historical, natural-forcing-only (hist-nat), SSP1–2.6, SSP2–4.5, SSP3–7.0, and SSP5–8.5. Historical attribution was evaluated over 1984–2014, and future projections were evaluated over 2020–2050. We retained nine CMIP6 models with complete daily coverage across all required experiments: ACCESS-ESM1-5, BCC-CSM2-MR, CanESM5, CNRM-CM6-1, GFDL-ESM4, IPSL-CM6A-LR, MIROC6, MRI-ESM2-0, and NorESM2-LM. For each CMIP6 experiment, daily near-surface air temperature (tas) fields were converted to degrees Celsius where necessary, calendar-normalized, and bilinearly interpolated to a ∼25 arc-minute grid^17^ using xESMF^65^. We then calculated ensemble means and inter-model standard deviations at each grid cell and time step, retaining these summaries for the historical, hist-nat, and SSP scenarios (Extended Data Fig. 6).

The primary endpoint for climate detection and attribution was continuous change in temperature-dependent transmission potential, quantified as model-derived R₀. We did not define detection solely as crossing a binary R₀ = 1 suitability threshold, because anthropogenic warming can amplify or suppress transmission potential within already suitable regions without causing a transition between unsuitable and suitable conditions. For each grid cell and year, daily mean temperature was propagated through the validated temperature-dependent R₀ framework to estimate annual transmission potential under each climate scenario. Following Aslan et al., areas lacking sufficient surface water or with low population density (<2 persons km⁻²) were masked from the analysis because sustained transmission is considered improbable in these regions^17^.

To isolate the anthropogenic temperature signal during the historical period, we compared transmission potential under the factual historical climate ensemble with transmission potential under the natural-forcing-only counterfactual ensemble. The anthropogenic contribution to transmission potential was calculated for each grid cell and year as ΔR₀ = R₀,_hist_ − R₀,_hist-nat_. Positive values indicate locations where anthropogenic warming increased temperature-dependent transmission potential, whereas negative values indicate thermal suppression. Proportional change was calculated relative to the natural-forcing baseline as ΔR₀/R₀,_hist-nat_.

Grid-cell estimates were aggregated to country-level boundaries from the Natural Earth database. For each country, we calculated the mean proportional change in R₀ and the 2.5th and 97.5th percentiles of grid-cell values to describe within-country spatial heterogeneity in transmission shifts. Population exposure was summarized using WorldPop 2015 population estimates^66^ to compare the number of people residing in areas where anthropogenic warming amplified versus suppressed transmission potential.

Using the same ensemble and spatial framework, we projected R₀ trajectories under SSP1–2.6, SSP2–4.5, SSP3–7.0, and SSP5–8.5 for 2020–2050. Future changes in transmission potential were calculated relative to the historical baseline period, 1984–2014. Because R₀ was calculated from ensemble climate summaries for computational efficiency, we used inter-model standard deviation in climate forcing to characterize variability among CMIP6 models. Historical changes are therefore described as ensemble-mean anthropogenic signals rather than statistically significant departures unless otherwise stated.

## Acknowledgements

We thank the Schistosomiasis Consortium for Operational Research and Evaluation (SCORE) investigators and the University of Georgia Research Foundation for generating the data used in this research, and the Schistosomiasis Resource Center for providing *Biomphalaria glabrata* snails and *Schistosoma mansoni* eggs. JRR was supported by the U.S. National Science Foundation (NSF; DEB-2109293, ITE-2333795, BCS-2307944, CISE-2435758), Notre Dame Poverty Initiative, Berthiaume Institute for Precision Health, and Frontiers Research Foundation. IA, AJC, GADL were supported by the NSF (ICER-2522282; DEB-2011179).

## Data availability statement

This study used multiple data sources, some of which are restricted, access-controlled, or not feasible to redistribute in raw form due to data-use limitations and/or file size. Restricted source data cannot be made publicly available by the authors. Processed or derived data generated by the authors will be made available where permissible. The analysis code will be made available in a public repository upon publication in a peer-reviewed journal.

## Author Contributions

MF and JRR conceptualized the study. MF led the investigation, data analysis, data visualization, and writing of the original draft. IA, CB, HB, AJC, GADL, KE, NAG, PH, KHN, MS, and JRR contributed to one or more of the following: data acquisition, methodological development, data analysis, and interpretation of results. All coauthors contributed to the writing and editing of the manuscript.

## Ethics statement

This study used secondary analyses of existing, de-identified human infection data from (1) the Schistosomiasis Consortium for Operational Research and Evaluation (SCORE) Gaining and Sustaining Control studies, obtained from ClinEpiDB in accordance with its data access and use policies, and (2) publicly available schistosomiasis prevalence data from the WHO Regional Office for Africa ESPEN portal. No new human participants were recruited, contacted, enrolled, sampled, or otherwise interacted with for the present study, and the authors did not have access to direct personal identifiers.

As reported for the ClinEpiDB SCORE dataset, written informed consent for participants in the Gaining and Sustaining Control studies was obtained from adults, including parents or legal guardians of participating children, and assent was obtained from children under 18 years of age, except in settings where village-level consent is the standard, in which case local requirements were met. The original research protocols were reviewed by the relevant human subjects committees and institutional review boards in each participating country and collaborating institution. The original SCORE studies were conducted under the ethical approvals and participant consent procedures applicable to those studies. The trials used in this study were registered with the International Standard Randomized Controlled Trial registry under ISRCT numbers 14849830 (Kenya Sustaining Control), 16755535 (Kenya Gaining Control), and 95819193 (Tanzania).

This study additionally included laboratory mesocosm experiments using Biomphalaria glabrata snails and Schistosoma mansoni eggs provided by the Schistosomiasis Resource Center. The authors did not house, maintain, infect, or euthanize vertebrate animals for this study. Handling of parasite material and freshwater snails was conducted in accordance with applicable institutional biosafety and containment procedures.

---

**Extended Data Figure 1.**
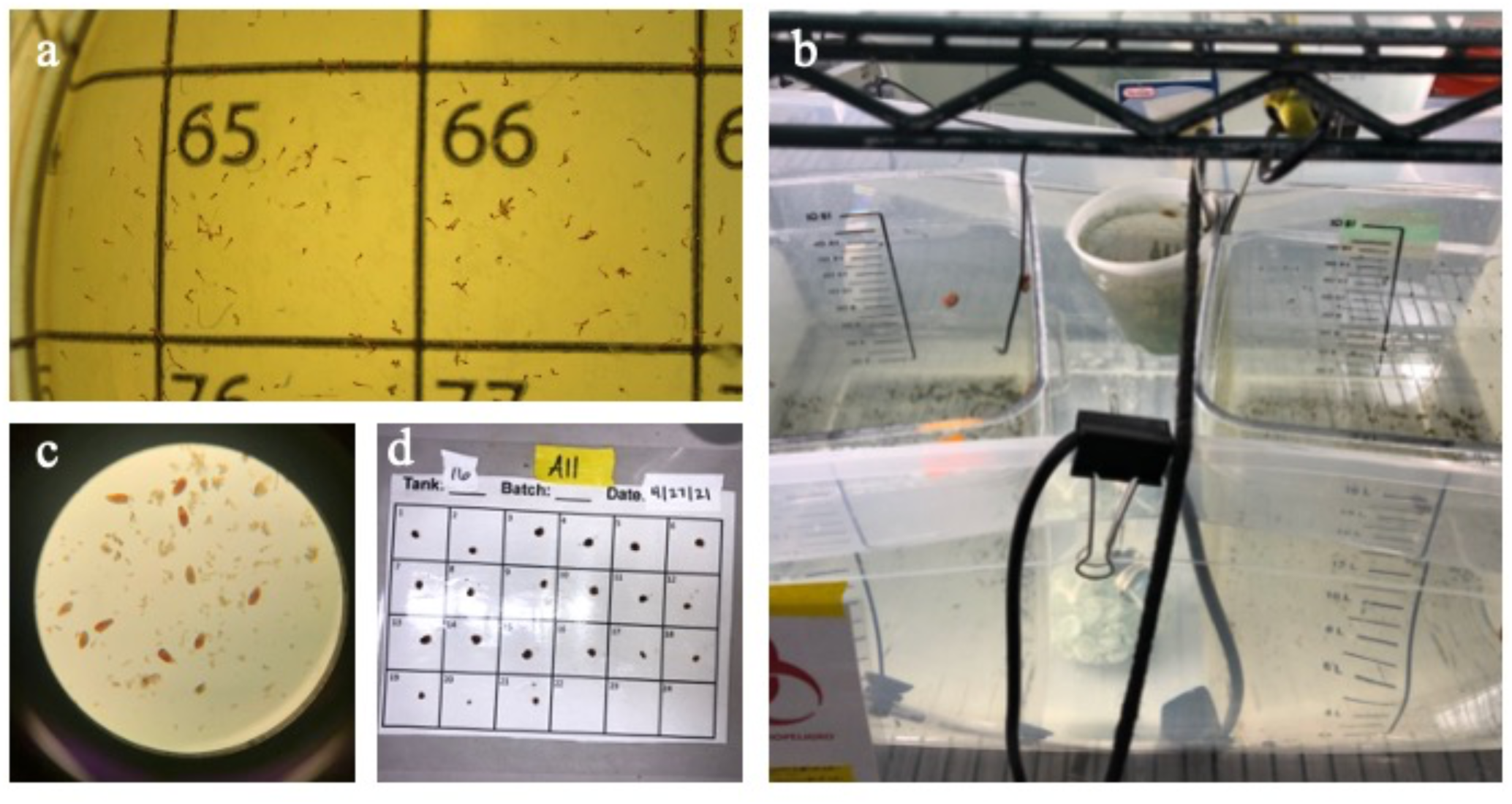
Experimental setup and data collection. (a) Grid-based subsampling of *Schistosoma mansoni* cercariae; (b) experimental mesocosms; (c) *S. mansoni* eggs; (d) *Biomphalaria glabrata* population counts.

**Extended Data Figure 2.**
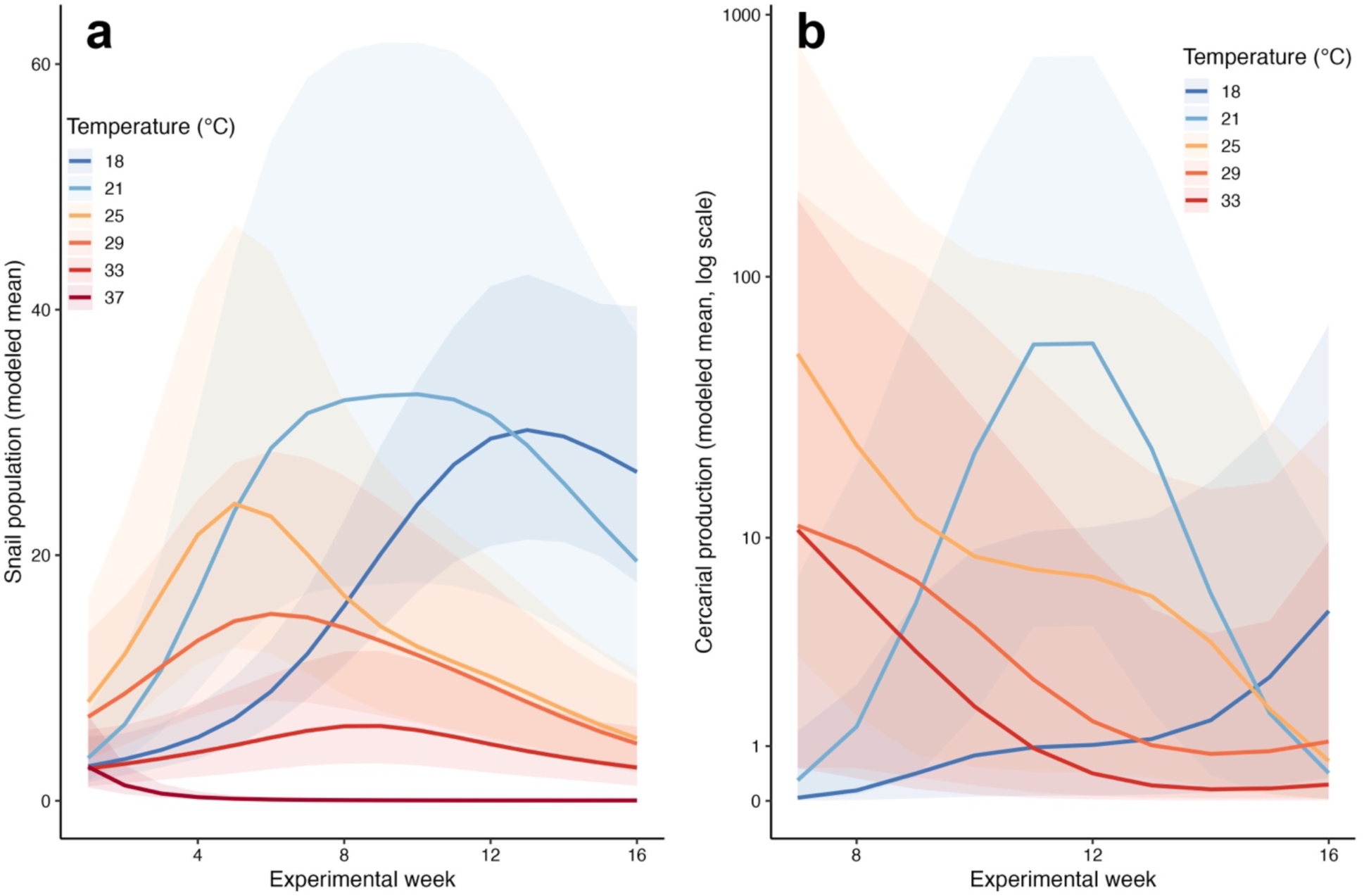
Temperature effects on snail populations and cercarial production in a 16-week mesocosm experiment. Dynamics of (a) *Biomphalaria glabrata* snail populations and (b) *Schistosoma mansoni* cercarial production (log scale) across a thermal gradient (18–37°C) over a two-temporal block 16-week mesocosm experiment. Lines represent modeled means from Generalized Additive Mixed Models (GAMMs) fitted to two experimental blocks (*n* =4 replicates per temperature per block); shaded ribbons indicate 95% confidence intervals.

**Extended Data Table 1.** Validation of thermal traits derived from experimental and mechanistic models: Comparison of thermal parameters for *Biomphalaria glabrata* snail populations and *Schistosoma mansoni* cercarial production. Values for the Experimental TPC represent the mean and 95% Bayesian credible intervals (CrI). Values for the Mechanistic TPC represent the mean and 95% confidence intervals (CI) from the model ensemble. T_opt_, T_min_, T_max_ represent the thermal optimum, lower limit, and upper limit, respectively, and were calculated using a 5% performance threshold. Note all uncertainty bands overlap with the exception of one endpoint. The mechanistic model underestimates upper thermal limit, resulting in conservative transmission boundaries.

| Response variable | Parameter (°C) | Experimental TPC (CrI) | Mechanistic TPC (CI) | Uncertainty bands overlap |
| --- | --- | --- | --- | --- |
| Snail population | $T_{opt}$ | 23.36 [21.90, 24.90] | 23.33 [22.27, 24.05] | Yes |
| | $T_{min}$ | 11.98 [10.50, 14.60] | 14.73 [12.29, 16.29] | Yes |
| | $T_{max}$ | 36.17 [35.60, 36.70] | 31.93 [30.20, 33.70] | No |
| Cercaria produced | $T_{opt}$ | 25.80 [23.60, 28.40] | 25.64 [20.34, 30.27] | Yes |
| | $T_{min}$ | 15.27 [14.00, 16.50] | 17.40 [11.64, 21.93] | Yes |
| | $T_{max}$ | 33.73 [33.1, 34.8] | 33.89 [24.82, 40.00] | Yes |

**Extended Data Figure 3.**
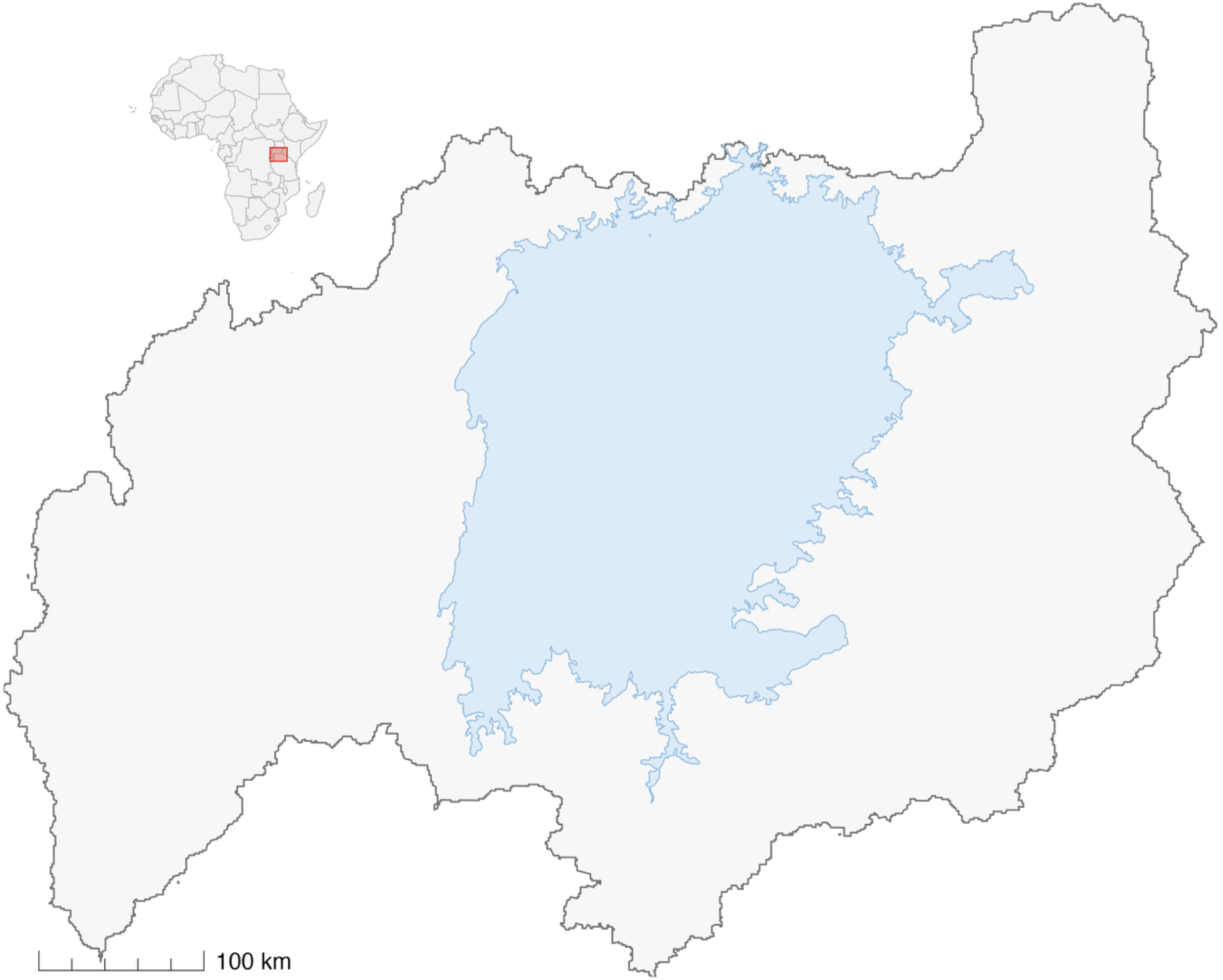
Map of the Lake Victoria Basin. The Lake Victoria Basin boundary is shown in grey, with Lake Victoria highlighted in blue. The epidemiological analysis included SCORE communities located within 10 km of the Lake Victoria shoreline; individual community locations are not shown to avoid disclosing potentially sensitive small-area human health-data locations. The red inset shows the location of the Lake Victoria Basin within Africa.

**Extended Data Table 2.** Longitudinal attribution model results within the Lake Victoria Basin. Fixed effects from a beta-binomial GLMM predicting *S. mansoni* prevalence among school-aged children (2011–2015). The model uses a Mundlak specification, separating climate exposures into within-community interannual deviations (Anomaly) and between-community long-term means (Baseline). Variance Inflation Factors (VIF) for all predictors were < 10.

| Predictor type | Predictor | Estimate<br>(log-odds) | Standard<br>error | P-value |
| --- | --- | --- | --- | --- |
| Climate variables | Temperature anomaly<br>(within-community) | 0.143 | 0.050 | 0.004 |
|  | Precipitation anomaly<br>(within-community) | 0.072 | 0.040 | 0.075 |
|  | Temperature baseline<br>(between-community) | -0.008 | 0.094 | 0.930 |
|  | Precipitation baseline<br>(between-community) | 0.292 | 0.203 | 0.151 |
| Intervention and geography<br>variables | Distance to lake | -0.522 | 0.058 | < 0.001 |
|  | Use of surface water | 0.544 | 0.216 | 0.012 |
| Study design variables | Treatment round 2 | 0.098 | 0.063 | 0.119 |
|  | Treatment round 3 | -0.637 | 0.071 | < 0.001 |
|  | Treatment round 4 | -1.053 | 0.072 | < 0.001 |
|  | Treatment round 5 | -1.240 | 0.113 | < 0.001 |
|  | Study arm effects | (Included) |  |  |

**Extended Data Table 3.** Performance comparison of geostatistical models for *S. mansoni* prevalence. Predictive accuracy was evaluated for seven zero-inflated beta-binomial models using 5-fold cross-validation. ‘Mean AUC’ represents the average Area Under the ROC Curve across folds. ΔAUC is the absolute improvement in predictive power compared to the Baseline model (AUC = 0.794). ‘Relative AUC gain’ quantifies the improvement of each model relative to the standard ‘Climate-Only’ reference (Baseline + T_min_ + T_max_).

| Model | Mean AUC | ΔAUC | Relative AUC gain |
| --- | --- | --- | --- |
| Baseline + $R_0$ | 0.849 | 0.0543 | 225% |
| Baseline + experimental population TPC | 0.849 | 0.0543 | 225% |
| Baseline + mechanistic cercariae TPC | 0.838 | 0.0437 | 162% |
| Baseline + mechanistic population TPC | 0.832 | 0.0378 | 126% |
| Baseline + experimental cercariae TPC | 0.822 | 0.0275 | 65% |
| Baseline + $T_{\min}$ + $T_{\max}$ | 0.811 | 0.0167 | — |
| Baseline | 0.794 | — | — |

**Extended Data Table 4.** Predictor importance for the continental geospatial model. Mean permutation importance for all covariates included in the final mechanistic zero-inflated beta-binomial GLMM predicting *S. mansoni* prevalence. Metrics were averaged across 5-fold cross-validation. Variables are sorted by descending relative importance.

| Predictor | Mean permutation importance |
| --- | --- |
| MDA coverage | 0.162 |
| GDP | 0.131 |
| R<sub>0</sub> (at T<sub>min</sub>) | 0.119 |
| Elevation | 0.105 |
| Water access | 0.0597 |
| Distance to lake | 0.0432 |
| R<sub>0</sub> (at T<sub>max</sub>) | 0.0138 |
| Sanitation | 0.0115 |
| Soil pH | 0.00530 |
| Urbanization | 0.000472 |
| Precipitation | -0.000160 |
| Infant mortality | -0.000212 |
| Mean soil moisture | -0.000731 |
| UN adjusted population | -0.000923 |
| Human modification of environment | -0.00646 |

**Extended Data Figure 4.**
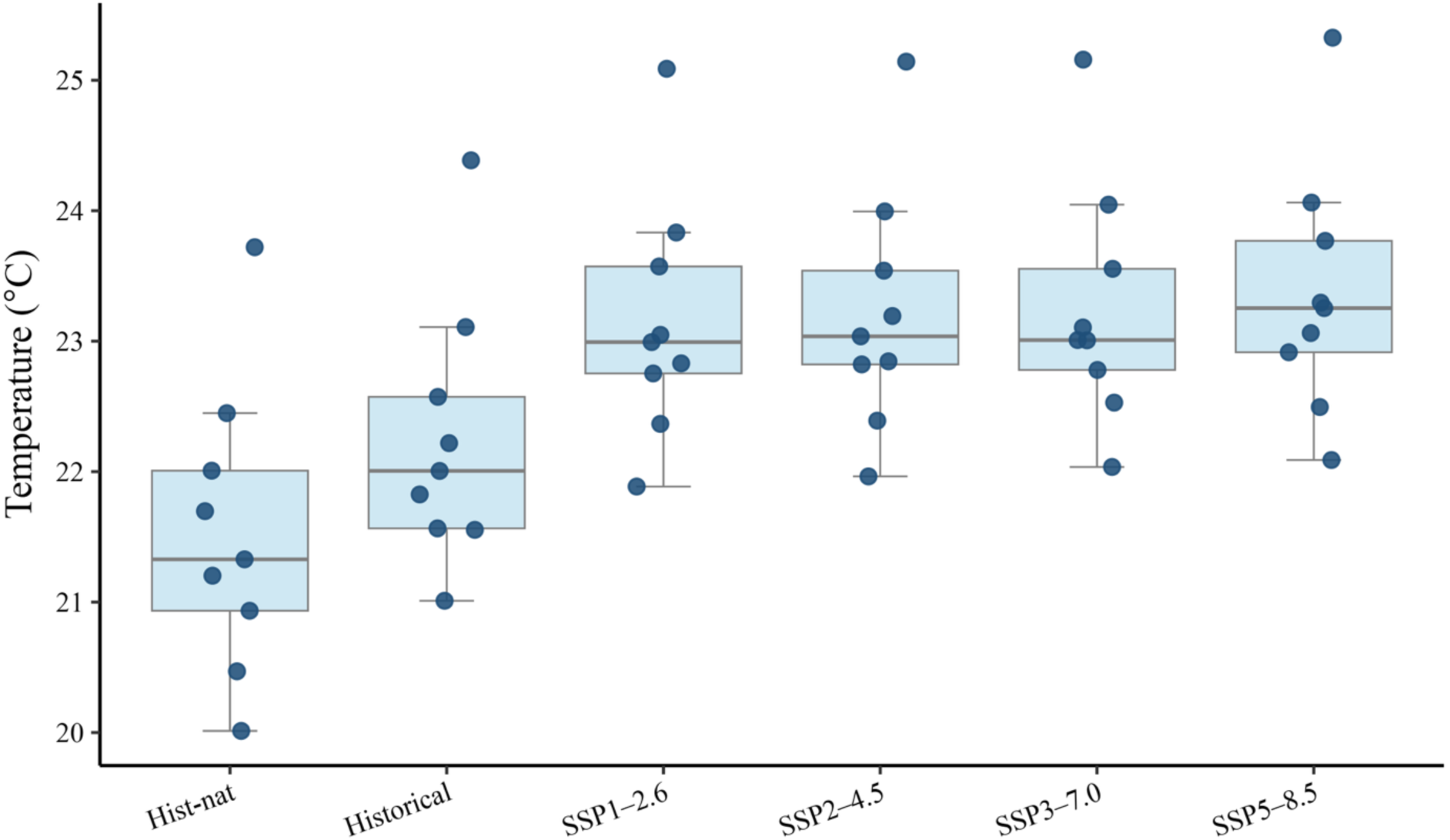
Inter-model variability in continental climate forcing across historical and future scenarios. The distribution of mean annual near-surface temperatures (tas) across Africa for the 9 Global Climate Models (GCMs). Boxes show the interquartile range, horizontal lines show the median, whiskers show the range across models, and points represent individual CMIP6 models. Each point represents a single GCM’s mean over a 31-year window: 1984–2014 for the natural-only counterfactual (hist-nat) and factual (historical) scenarios, and 2020–2050 for the four Shared Socioeconomic Pathway (SSP) future projections.

**Extended Data Figure 5.**
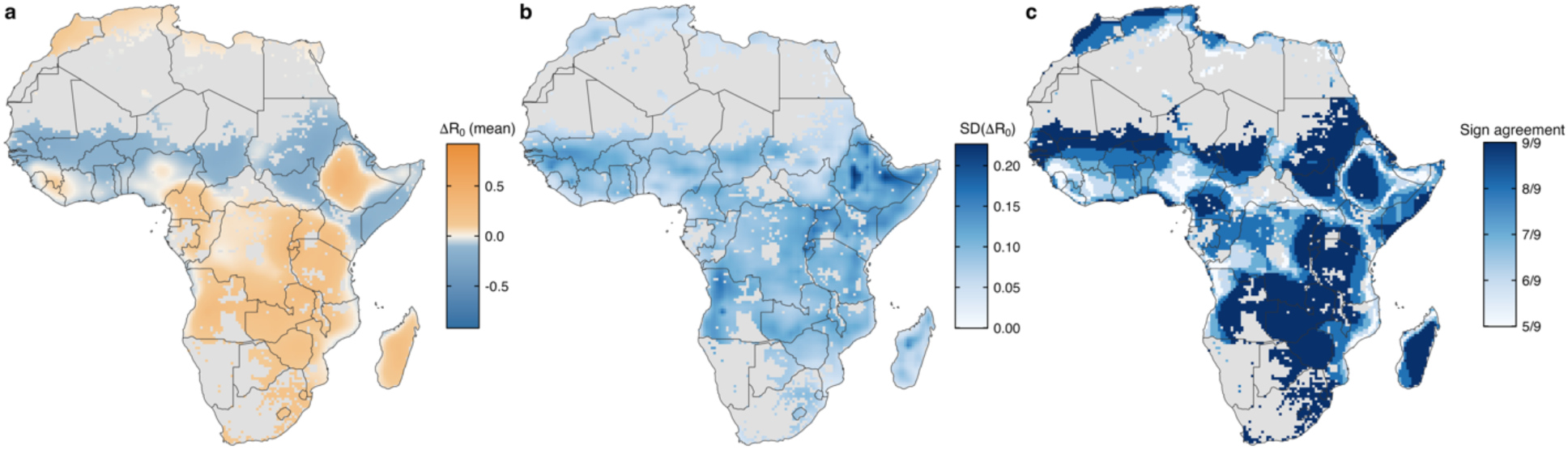
CMIP6 ensemble uncertainty in anthropogenic-warming-driven changes in schistosomiasis transmission potential. Maps show anthropogenic warming changes in temperature-dependent transmission potential, calculated as ΔR₀ = R₀,_historical_ − R₀,_hist-nat_ and averaged over 1984–2014. (a) The ensemble-mean ΔR₀ across the 9 CMIP6 models. Positive ΔR₀ values indicate increased transmission potential under anthropogenic warming, while negative values indicate thermal suppression. Grey areas indicate cells excluded from the transmission mask. (b) The inter-model standard deviation in ΔR₀. (c) Agreement in the sign of ΔR₀ across models, shown as the number of models sharing the majority sign at each grid cell.

**Extended Data Figure 6.**
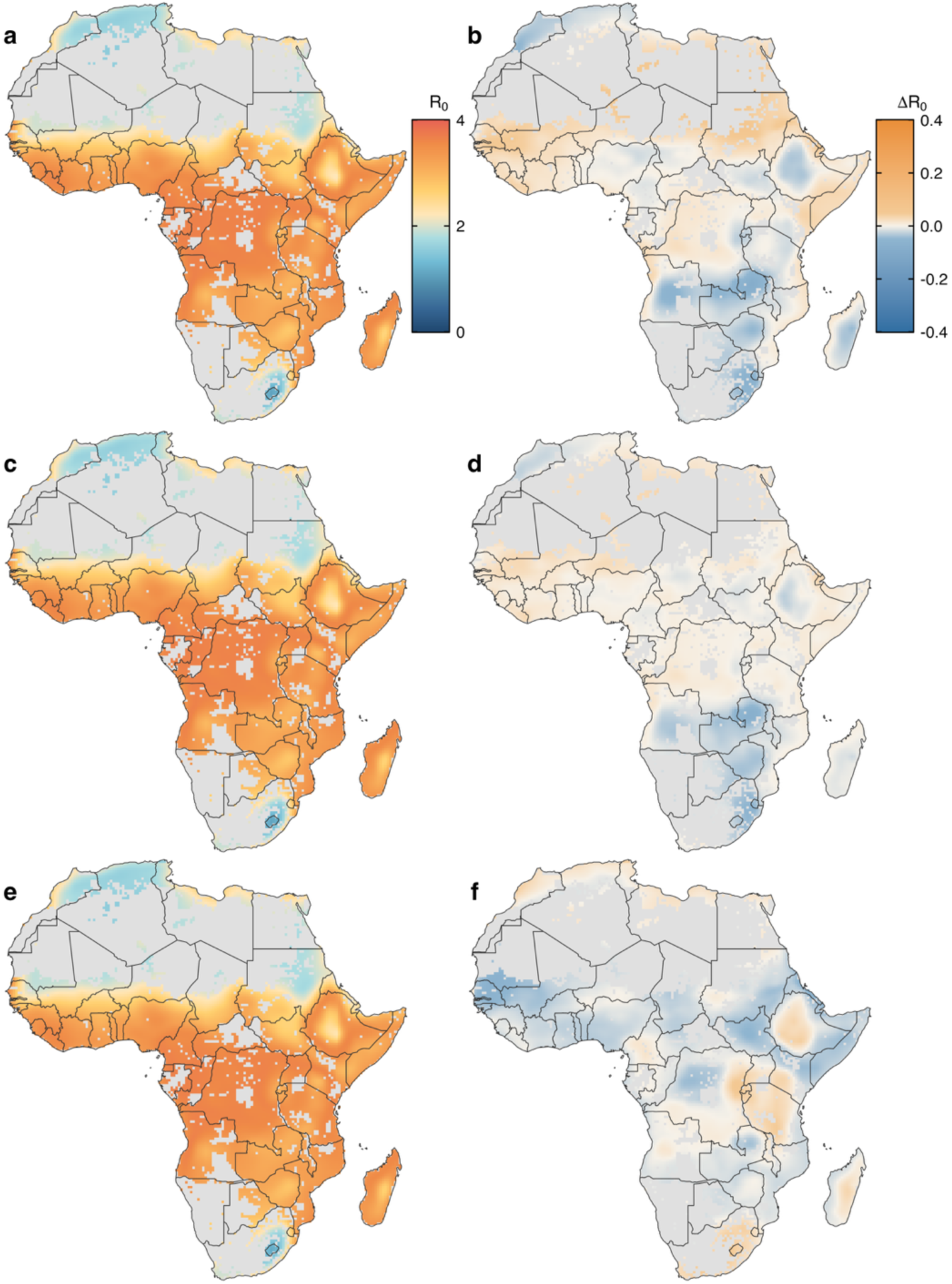
Projected transmission potential under alternative SSP scenarios and deviations from SSP3-7.0. Maps show multi-model ensemble projections of annual schistosomiasis transmission potential (R₀) across continental Africa under (a) SSP1–2.6, (c) SSP2–4.5, and (e) SSP5–8.5. Corresponding difference maps quantify deviations relative to the SSP3–7.0 scenario shown in the main text: (b) Δ R₀= SSP1–2.6 minus SSP3–7.0, (d) ΔR₀ = SSP2–4.5 minus SSP3–7.0, (f) ΔR₀ = SSP5–8.5 minus SSP3–7.0. R₀ was generated by driving the validated mechanistic transmission model with daily near-surface air temperature from a nine-model CMIP6 ensemble.

**Supplementary Table 1.** Predictor variables used in the continental geostatistical models. Temporal resolution, spatial resolution or spatial unit, and data source for predictor variables included in models of *S. mansoni* prevalence across Africa.

| Predictor | Temporal resolution | Spatial resolution | Source |
| --- | --- | --- | --- |
| Water supply | Annual | Implementation unit | ESPEN |
| Sanitation | Annual | Implementation unit | ESPEN |
| Mean MDA coverage | Current and prior survey year | Implementation unit | World Health Organization |
| Mean annual minimum temperature | Monthly climatology, aggregated to annual mean | 2.5 arc-minutes | WorldClim v2.1 |
| Mean annual maximum temperature | Monthly climatology, aggregated to annual mean | 2.5 arc-minutes | WorldClim v2.1 |
| Mean annual precipitation | Monthly climatology, aggregated to annual total | 2.5 arc-minutes | WorldClim v2.1 |
| Elevation | Static | 2.5 arc-minutes | WorldClim v2.1 |
| Mean soil pH | Static soil property | 5 arc-minutes | ISRIC-WISE v1.2 |
| Gross domestic product, purchasing power parity | Annual | 5 arc-minutes | Kummu et al., 2018; Dryad data set |
| Human modification of terrestrial systems | Static; circa 2016 | 1 km<sup>2</sup> | Kennedy et al., 2020; NASA SEDAC Global Human Modification data set v1.00 |
| Mean soil moisture | Annual | 0.25° | ESA CCI Soil Moisture |
| Urbanization | 2015 or nearest available year | Urban-center polygons derived from 1 × 1 km grid | GHSL Urban Centre Database |
| UN-adjusted population | Annual | 1 km | WorldPop |
| Infant mortality | 2015 | National/subnational units | CIESIN/NASA SEDAC |
| Distance to nearest lake | Static | Vector hydrography | FAO AQUASTAT |
| Distance to nearest river | Static | Vector hydrography | FAO AQUASTAT |

**Supplementary Table 2.** Spatial analysis for continental geostatistical models. Model performance after spatial checkerboard partitioning using approximately 100-km blocks. Models are ordered by spatially partitioned AUC. AIC support groups summarize overlap in model support using a ΔAIC threshold of 2. Models sharing support-group labels were treated as having overlapping AIC support under the spatial partitioning analysis.

| Model | Spatially partitioned AUC | AIC | ΔAIC from lowest AIC | AIC support group |
| --- | --- | --- | --- | --- |
| Baseline + experimental population TPC | 0.858 | 34,762.21 | 8.48 | B |
| Baseline + experimental cercariae TPC | 0.855 | 34,785.19 | 31.46 | D |
| Baseline + mechanistic cercariae TPC | 0.830 | 34,765.05 | 11.32 | BC |
| Baseline + $R_0$ | 0.829 | 34,763.07 | 9.34 | B |
| Baseline + mechanistic population TPC | 0.821 | 34,753.73 | 0.00 | A |
| Baseline + $T_{\min}$ + $T_{\max}$ | 0.807 | 34,765.50 | 11.77 | C |
| Baseline | 0.770 | 34,851.43 | 97.70 | E |

